# Diverse ancestry GWAS for advanced age-related macular degeneration in TOPMed-imputed and Ophthalmologically-confirmed 16,108 cases and 18,038 controls

**DOI:** 10.1101/2024.11.08.24316962

**Authors:** Mathias Gorski, Michelle Grunin, Janina M. Herold, Benedikt Fröhlich, Merle Behr, Nicholas Wheeler, William S. Bush, Yeunjoo E. Song, Xiaofeng Zhu, Susan H. Blanton, Margaret A. Pericak-Vance, Iris M. Heid, Jonathan L. Haines, International Age-related Macular Degeneration Genomics Consortium

## Abstract

Age-related macular degeneration (AMD) is a leading cause of blindness with $344 billion dollars global costs. In 2016, the International Age-related Macular Degeneration Genomics Consortium devised genomic data on ∼50,000 individuals (IAMDGC 1.0) and identified 52 variants across 34 loci associated with advanced AMD in European ancestry. We have now analyzed a more densely imputed version (IAMDGC 2.0) and performed cross-ancestry GWAS in 16,108 advanced AMD cases and 18,038 AMD-free controls. This identified 28 loci at P<5×10^−8^, including two additional AMD loci compared to IAMDGC 1.0 (*SERPINA1* and *CPN1*). Fine-mapping supported one ancestry-shared signal around *HTRA1/ARMS2* and nine signals around *CFH* without African ancestry contribution. The 52-variant genetic risk score with and the 44-variant score without *CFH*-variants predicted advanced AMD not only in EUR, but also in AFR and ASN (AUC=0.80/0.75, 0.65/0.64, 0.80/0.79, respectively). Our results indicate that the genetic underpinning of advanced AMD is mostly shared between ancestries.

---

Advanced AMD is characterized by the growth of abnormal pathological blood vessels into the retina, known as neovascular AMD (nvAMD), or the destruction of cells in specific areas as the retinal pigment epithelium ceases to function, known as geographic atrophy (GA). At present, the only available therapies target nvAMD to slow the growth of new blood vessels into the retina^1^ or GA via cell therapy to try replacing dead RPE cells. However, a permanent cure and a full understanding of the pathogenesis of AMD is lacking.

In 2016, the International Age-related Macular Degeneration (AMD) Genomics Consortium (IAMDGC) devised a large 1000 Genomes imputed dataset including >50,000 individuals of multiple ancestries (IAMDGC1.0, hg37) and their GWAS of 16,144 advanced AMD cases and 17,832 AMD-free controls of European ancestry (EUR) identified 52 independent variants across 34 loci. The IAMDGC 1.0 dataset and its GWAS summary statistics in EUR are have been an important resource for many meta-analyses^2–4^, Mendelian Randomization studies^5–8^ and comparisons of advanced AMD with related traits^9–12^. By this, IAMDGC 1.0 has fostered the understanding of genetic and non-genetic causes for advanced AMD. The IAMDGC dataset had to be updated due to study termination of a subset of individuals and the denser imputation reference panel from TOPMed and hg38 becoming state-of-the-art. Importantly, the previous GWAS did not include the African American (AFR), Asian American (ASN) or other ancestry individuals recruited by IAMDGC. Diverse ancestry GWAS are sparse for AMD. One GWAS meta-analyzing the multi-ancestry Million Veterans Project (MVP) and the previous EUR-based IAMDGC GWAS results identified the first loci for advanced AMD in AFR and Hispanic Americans^4^. Since only 9% of the MVP dataset are women, it remains important to evaluate the contribution of both men and women in the cross-ancestry genetic underpinning of AMD. Therefore, we set out to re-impute the IAMDGC dataset using the TOPMed reference panel based on hg38 (IAMDGC 2.0), to re-analyze the EUR-part of the dataset, and to conduct new GWAS in AFR, ASN, and other ancestry individuals.

We utilized the centralized genotyped data from IAMDGC as described previously^13^ removing individuals from the Beaver Dam Eye Study (study termination) or with Whole Genome Amplification (potential disruption of imputation^14^, **Methods**). This reduced the dataset from 52,189 to 44,401 individuals. After TOPMed-imputation and quality control (**Methods**), we compared the TOPMed-imputed dataset (IAMDGC 2.0) with the previous 1000G-imputed dataset (IAMDGC 1.0). Among the 292,027,721 initially imputed autosomal variants (**Supplementary Table 1**), the number of high-quality imputed variants (RSQ≥0.3 for MAF≥1%, RSQ≥0.8 for MAF<1%) is now 33,661,604 versus 11,602,234 previously (**Table 1**). Restricting analyses to genetic variants that were robustly analyzable in at least one ancestry (minor allele count, MAC, ≥20^15^, **Methods**) yielded 19,299,720 well-imputed autosomal variants compared to 10,231,250 such variants previously (for rare variants,

**Table 1:** Analyzed individuals and well-imputed autosomal genetic variants. We imputed the 46,601 individuals with centrally measured genotypes with the TOPMed references data (Biocatalyst imputation server, hg38) comprising the IAMDGC 2.0 dataset. We here show the number of individuals by case-control status and overall, of well-imputed autosomal variants, and of well-imputed (MAF≤0.01 and RSQ≥0.8 or MAF>0.01 and RSQ≥0.3) and well-imputed and analyzable variants (additionally requiring MAC≥20 per ancestry) in the IAMDGC 2.0 dataset. Shown are absolute numbers by ancestry (EUR=European, AFR=African, ASN=Asian and OTH=other ancestry), in the cross-ancestry dataset of IAMDGC 2.0 and for IAMDGC 1.0^13^.

|  | IAMDGC 2.0 |  |  |  |  |  |
| --- | --- | --- | --- | --- | --- | --- |
|  | EUR | AFR | ASN | OTH | Cross-ancestry | IAMDGC 1.0 |
| Analyzed Individuals |  |  |  |  |  |  |
| Overall | 32,339 | 407 | 529 | 871 | 34,146 | 33,976 |
| Cases | 15,616 | 50 | 207 | 235 | 16,108 | 16,144 |
| Controls | 16,723 | 357 | 322 | 636 | 18,038 | 17,832 |
| Well imputed genetic variants |  |  |  |  |  |  |
| Overall | 24,453,272 | 17,779,590 | 8,491,777 | 18,824,278 | 33,661,608 | 11,602,234 |
| MAF≥1% | 8,890,336 | 15,486,439 | 8,027,043 | 13,347,013 | 25,447,456 | 8,880,799 |
| MAF<1% | 15,562,936 | 2,293,151 | 464,734 | 5,477,265 | 8,214,152 | 2,721,435 |
| Well imputed and analyzable in at least one ancestry (MAC≥20) |  |  |  |  |  |  |
| Overall | 16,251,760 | 10,984,978 | 6,998,030 | 12,380,946 | 19,299,720 | 10,231,250 |
| MAF≥1% | 8,887,558 | 10,984,978 | 6,998,030 | 12,380,946 | 13,474,841 | 8,862,912 |
| MAF<1% | 7,364,202 | 0 | 0 | 0 | 5,824,879 | 1,368,338 |

Among the 9,485,579 variants in both IAMDGC 2.0 and IAMDGC 1.0 imputation, there were 6,722,468 common, 2,323,652 less frequent and 439,459 rare analyzable variants.

MAF<1%: 5,824,879 versus 1,368,338 previously; **Table 1**). Thus, IAMDGC 2.0 versus IAMDGC 1.0 improved coverage of autosomal genetic variants for GWAS by nearly 100% (400% for rare variants). Imputation quality was similar for variants with MAF≥1% (median RSQ=0.97 versus 0.94, respectively), but substantially improved for rare variants (median RSQ=0.97 versus 0.88, respectively; among 9,485,579 well-imputed variants analyzable in both IAMDGC 2.0 and 1.0, including 439,459 rare variants). This underscores the gain in genome coverage and imputation quality for GWAS, particularly for rare variants.

We conducted ancestry-specific GWAS (cases/controls=15,616/16,723 EUR, 50/357 AFR, 207/322 ASN, and 235/636 Other; **Table 1**) using Firth test logistic regression implemented in regenie^16^ adjusted for two ancestry-specific PCs and their meta-analysis in a total of 16,108 advanced AMD cases and 18,038 AMD-free controls (**Methods, Table 1**). Despite adding non-EUR ancestry and related individuals, this cross-ancestry IAMDGC 2.0 GWAS resulted in a similar sample size as the IAMDGC 1.0 GWAS due to removing the Beaver Dam Eye Study and individuals with Whole Genome Amplification (i.e. power slightly lower now due to related individuals). Per-ancestry GWAS was GC-corrected where applicable (lambda =1.11, 0.99, 0.98, and 1.08 in EUR, AFR, ASN, Other, respectively; **Supplementary Figure 1**), resulting in lambda=0.99 in the cross-ancestry GWAS meta-analysis (**Figure 1A&B**). When querying the 13,474,841 variants with MAF≥1% at P<5×10^−8^ in cross-ancestry GWAS results, we found 28 loci (±500kb around each lead variant; merging overlapping loci; **Methods**): 26 of the 34 loci previously identified and two new loci compared to IAMDGC 1.0 (**Supplementary Table 2**). The 26 locus lead variants identified here were mostly the same or correlated (r≥0.8) to the lead variants from Fritsche et al. (except for three loci, **Supplementary Figure 2A-F**). The other 8 of the 34 previous AMD loci were not identified genome-wide (likely due to lower power), but detectable at P<1×10^−4^ (**Supplementary Table 2;** same/correlated lead variants here as before except for 4 loci, **Supplementary Figure 2G-N**). Extending GWAS to rare variants (adding 5.3 million variants with MAF<1%) did not detect additional loci at P<1×10^−9^. The 36 AMD locus lead variants showed similar ORs and standard errors cross-ancestry versus EUR-only (IAMDGC 2.0) and similar EUR-based ORs, standard errors, and risk-increasing allele frequencies (RAF) in controls (proxy for population) in IAMDGC 2.0 versus IAMDGC 1.0 (**Supplementary Figure 3; Supplementary Data 1**). Sensitivity analyses with alternative adjustments showed similar lambda and same ORs (**Supplementary Data 2**). Overall, the cross-ancestry GWAS indicated that the 34 known AMD loci generalize across ancestries and identified two additional loci in this dataset.

**Table 2:** The 52-variant GRS predicts advanced AMD across ancestries. We built the GRS based on 52 variants identified in Fritsche et al.^13^, and their beta-estimates as weights. The weighted GRS was calculated in the IAMDGC 2.0 data for EUR, AFR, ASN, and Other (n_cases_/n_controls_ = 15,616/16,723, 50/357, 207/322, 235/636, respectively). We also used different subsets of variants to calculate a GRS: (i) a 42-variant GRS including only variants analyzable in each ancestry (“shared variants”; risk allele count, RAC, ≥20 and 2^*^n – RAC ≥20 in EUR, AFR, ASN, and Other), (ii) a 3-variant GRS using only variants in the *CFH* locus that were analyzable in each ancestry and (iii) a 44-variant GRS excluding *CFH* variants. For each ancestry and each GRS, we applied a logistic regression model (advanced AMD ∼ GRS, 2 PCs). Shown are Odds Ratios with 95% CIs and association p-values. Also shown is the area-under-the-curve (AUC) for the model’s ability to separate cases from controls using predicted probabilities from the respective logistic regression models.

| GRS | EUR | AFR | ASN | OTHER |
| --- | --- | --- | --- | --- |
| **GRS_52 (52)** |  |  |  |  |
| OR [95% CI] | 1.46 [1.45, 1.48] | 1.21 [1.08, 1.34] | 1.32 [1.22, 1.43] | 1.34 [1.26, 1.42] |
| p-value | $<1.0 \times 10^{-320}$ | $1.0 \times 10^{-3}$ | $1.9 \times 10^{-12}$ | $2.2 \times 10^{-22}$ |
| AUC | 0.80 | 0.65 | 0.80 | 0.78 |
| **GRS shared (42)** |  |  |  |  |
| OR [95% CI] | 1.27 [1.26, 1.27] | 1.12 [1.05, 1.20] | 1.19 [1.13, 1.25] | 1.21 [1.17, 1.26] |
| p-value | $<1.0 \times 10^{-320}$ | $1.1 \times 10^{-3}$ | $2.3 \times 10^{-12}$ | $3.8 \times 10^{-23}$ |
| AUC | 0.79 | 0.63 | 0.80 | 0.78 |
| **GRS_CFH_only (3)** |  |  |  |  |
| OR [95% CI] | 1.45 [1.43, 1.47] | 1.11 [0.90, 1.36] | 1.32 [1.13, 1.55] | 1.46 [1.33, 1.62] |
| p-value | $<1.0 \times 10^{-320}$ | 0.34 | $4.8 \times 10^{-04}$ | $2.6 \times 10^{-14}$ |
| AUC | 0.68 | 0.53 | 0.74 | 0.74 |
| **GRS no CFH (44)** |  |  |  |  |
| OR [95% CI] | 1.34 [1.33, 1.35] | 1.17 [1.07, 1.29] | 1.24 [1.22, 1.43] | 1.23 [1.16, 1.29] |
| p-value | $<1.0 \times 10^{-320}$ | $7.0 \times 10^{-4}$ | $1.2 \times 10^{-10}$ | $3.8 \times 10^{-14}$ |
| AUC | 0.75 | 0.64 | 0.79 | 0.73 |

**Figure 1:**
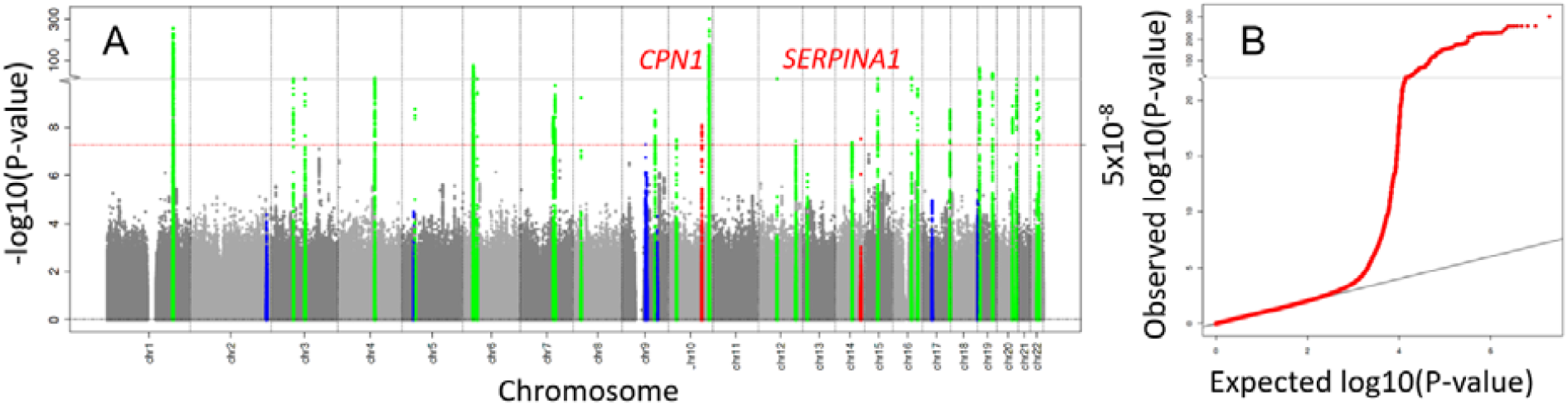
Cross-ancestry GWAS in IAMDGC 2.0 identifies 26 known AMD loci and two additional loci. We conducted ancestry-specific GWAS in the IAMDGC 2.0 data (Firth test based logistic regression adjusted for ancestry-specific PCs, using regenie^16^) and cross-ancestry GWAS meta-analysis (16,108 advanced AMD cases, 18,038 controls; ∼20 million autosomal TOPMed-imputed variants, hg38). Shown are (**A**) P-values for association, highlighting the 26 known loci identified at P<5×10^−8^ in green, the two loci new in IAMDGC 2.0 at P<5×10^−8^ versus IAMDGC 1.0 in red (near *CPN1*, chr10; *SERPINA1*, chr14), and the other 8 of the 34 known AMD identified at P<1×10^−4^ in blue. Also shown is (**B**) the QQ plot illustrating observed versus expected p-values (lambda=0.99).

**Figure 2:**
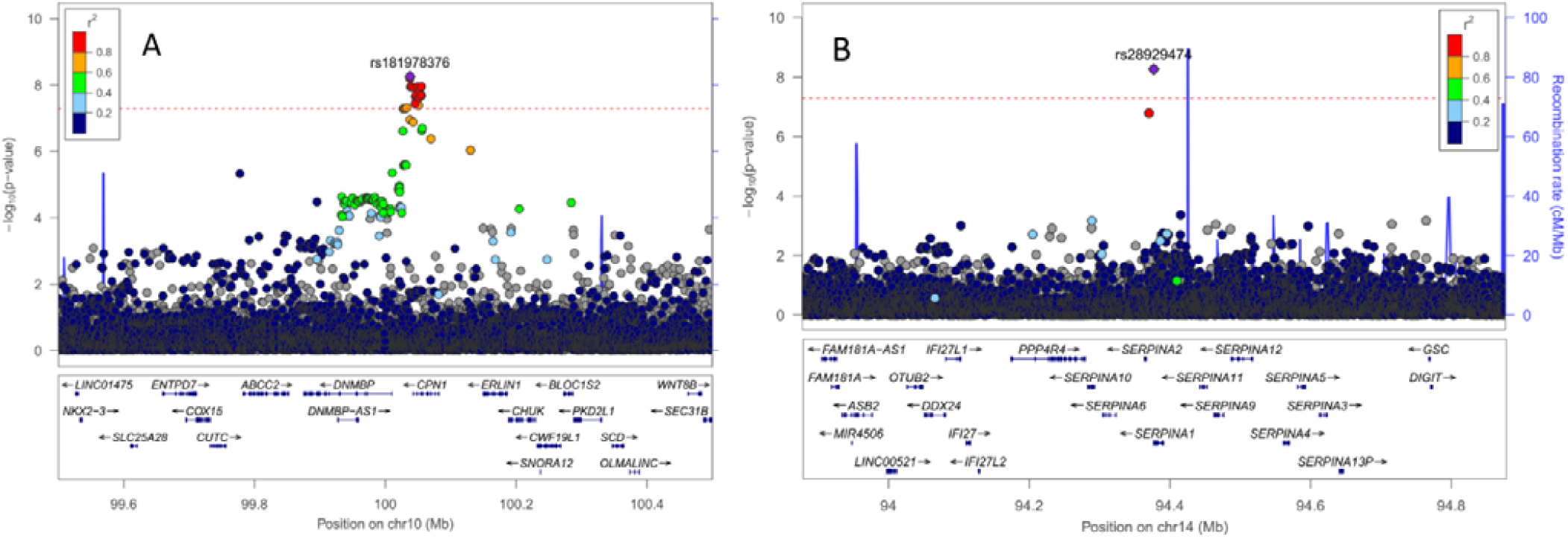
Two additional loci in the IAMDGC 2.0 cross-ancestry GWAS not identified in the previous IAMDGC 1.0 GWAS. We compared the association signals of the two loci identified newly in IAMDGC 2.0 (16,108 advanced AMD cases, 18,038 controls). Shown are regional association P-values in (**A**) the *CPN1* locus and (**B**) the *SERPINA1* locus. The variant with the smallest P-value (lead variant) is highlighted as violet diamond.

**Figure 3:**
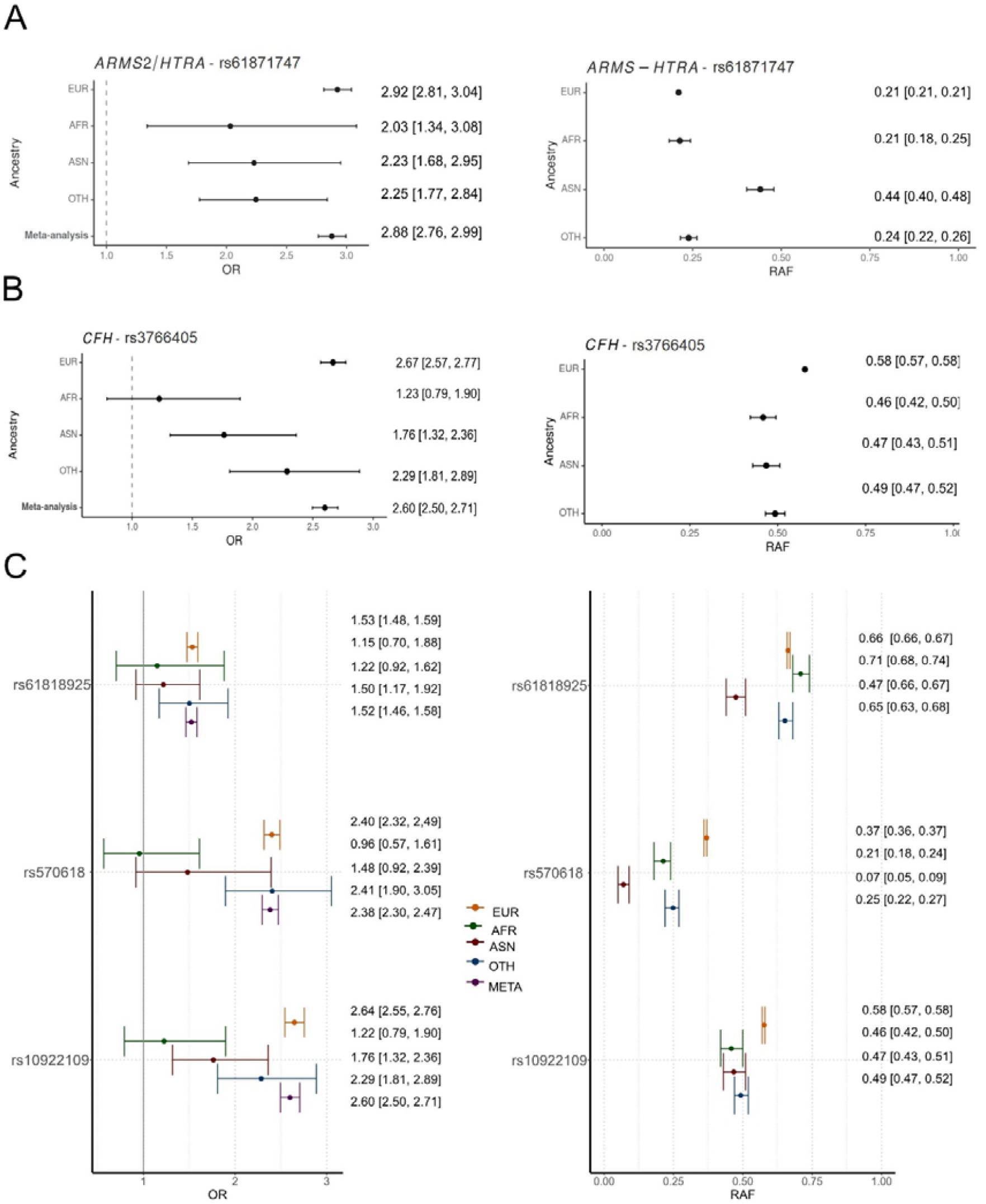
Ancestry-specific association for variants in *ARM2-HTRA1* and *CFH* locus. We compared ancestry-specific association results for variants in the *ARMS2-HTRA* and *CFH* loci (European, EUR, African, AFR, Asian, ASN, and other ancestries; cases/controls = 15,616/16,723, 50/357, 207/322, 235/636 respectively). Risk allele frequency (RAF) in controls as proxy for population frequency was calculated in each ancestry separately. Shown are (left) ancestry-specific and cross-ancestry meta-analyzed odds ratios and 95%-CIs and (right) RAF of **(A)** the lead variant rs61871747 in the *ARMS2-HTRA1*, (**B**) the lead variant rs3766405 in the *CFH* locus, and (**C**) for three of the eight independent *CFH* variants identified previously^13^ that were analyzable in each ancestry (MAC≥20, rs61818925, rs570618 and rs10922109).

The two additional loci identified in the IAMDGC 2.0 cross-ancestry GWAS compared to the EUR-only IAMDGC 1.0 resided around *CPN1* and *SERPINA* (P=8.32×10^−9^, OR=1.28, RAF=0.045 and P=2.95×10^−8^, OR=0.1.54, EAF=0.016; respectively, **Figure 2A&B**; previous P=2.55×10^−7^ and 1.50×10^−6^, respectively). Although new to the IAMDGC analysis, both have been recently reported: (i) the locus around *CPN1* was reported^3^ from a meta-analysis of two EUR-only datasets of this consortium: from 2016^13^ (IAMDGC 1.0, centralized genotyping) and an older dataset from 2013^17^ (federated genotyping) with largely overlapping individuals. (ii) The *SERPINA* locus was reported by Gorman et al.^4^ meta-analyzing MVP and IAMDGC 1.0 (total of 61,248 cases and 364,472 controls). The lead variant for *SERPINA* here, rs28929474, is highly correlated to the locus lead variant in Gorman et al. (rs112635299, r=0.99) and a missense variant predicted to change protein folding (VEP consequence score=7; Missense3D, UniProt Consortium 2023^18^. Thus, this IAMDGC 2.0 dataset more firmly identified the *CPN1* locus compared to Han et al. and the *SERPINA* locus now with ∼50% women compared to Gorman et al. prioritizing the variant that changes SERPINA1 protein structure.

The most prominent AMD loci in this cross-ancestry GWAS are, again, around *HTRA1/ARMS2* and *CFH*, and their large effects on AMD allow for cross-ancestry comparison. Their lead variants showed significant between-ancestry heterogeneity (P_het_=0.01 and 0.001; I^2^=70.8 and 84.3, respectively). We evaluated between-ancestry similarity or difference by examining the 95%-CIs of ORs: for the *HTRA1/ARMS2* locus lead variant, rs61871747, ancestry-specific 95%-CIs excluded unity and overlapped each other, which indicates no effect difference between ancestries (ORs for risk-increasing allele between 2.03 to 2.92; **Figure 3A**). For the *CFH* locus lead variant, rs3766405, 95%-CIs excluded unity in EUR, ASN and Other (OR for risk-increasing allele between 1.76 and 2.67), but not in AFR (P=0.36; OR [95%CI] = 1.23 [0.79, 1.90]; **Figure 3A**). The 95%-CIs of EUR and AFR or ASN did not overlap, which indicates between-ancestries differences. The observed lack of association in AFR could be indicative of truly no effect, but it could also result from lack of power in AFR (50 cases, 357 controls). However, the non-AFR OR is similar for the *CFH* variant and the *HTRA1/ARMS2* variant, OR∼2.0, and the *HTRA1/ARMS2* variant association was detectable in AFR at P<0.05. This suggests that we had power to detect a *CFH* variant effect as large as OR∼2.0. Ancestry-differential allele frequency, *f*, could result in lower genotype variance (var_G_ = 2^*^f^*^(1-f)) in AFR and thus comprised power. However, the genotype variance of the *CFH* lead variant was similar in EUR and AFR (var_G_=0.49 and 0.50 based on RAF=0.58 and 0.46, respectively; **Figure 3B**), which rules out lower power in AFR due to lower genotype variance. Differential effects across ancestries can also indicate that the studied variant is not causal. More likely causal might be the eight independent variants in the *CFH* locus identified by sequential forward selection previously^13^, of which three variants were analyzable in each ancestry (MAC≥20; rs10922109, rs570618, and rs61818925), but again none of these variants showed association in AFR (**Figure 3C**). Overall, our results suggest that, in individuals of AFR ancestry, there is no effect of *CFH* or a substantially lower effect than in EUR, which is in line with previous reports^4^. Our results also indicate a similar effect of *HTRA1*/*ARMS* across ancestries, while results from MVP^4^ suggested substantially lower effects in AFR compared to EUR.

Multi-ancestry fine-mapping can provide information whether the signals, which consist of many partly correlated variants, and not only the index variants, are ancestry-shared or ancestry-specific. We thus conducted multi-ancestry fine-mapping in the *HTRA1/ARMS2* and *CFH* loci for EUR and AFR individuals using MESusiE^19^ based on variants analyzable in both ancestries (**Methods**): (i) for *HTRA1/ARMS2*, we found one signal that was ancestry-shared (posterior inclusion probability of shared, EUR-specific, or AFR-specific: 86%, 14%, 0%, respectively; **Supplementary Data 3**). The 95% credible set contained ten variants with approximately equal probabilities to be the likely association-driving variant (all in/near *ARMS2*). (ii) For *CFH*, we found nine signals that were most likely EUR-specific (posterior inclusion probability: 25-39%, 61-76% or 0%, respectively; **Supplementary Data 3**). Since MESusiE requires variants to be analyzable in both EUR and AFR, which impedes analyses of less frequent variants due to the smaller AFR sample size, we repeated fine-mapping in EUR-only via SuSiE^20^ adding the less frequent variants analyzable variants in EUR (**Methods**). We found (i) the same number of signals in the *CFH* locus and (ii) now two signals in *HTRA1/ARMS2*, including one additional rather rare single-variant signal residing in *HTRA1* (rs13322423 with posterior inclusion probability=1, RAF=1.1% in EUR controls). Thus, our multi-ancestry fine-mapping results supported an ancestry-shared signal around *ARMS2* together with a rare variant in *HTRA1* that was not analyzable in AFR and little or no effect of *CFH* signals in AFR.

The 52 variants and their beta-estimates identified previously in IAMDGC 1.0 EUR-only^13^ are widely used for genetic risk score (GRS) analyses (e.g. Gorman et al.^4^, Grunin et al.^21^, Hesteerbeek et al.^22^, Yu et al.^23^). We were thus interested in whether the results for these 52 variants still hold in the updated IAMDGC 2.0 dataset and whether the 52-variant GRS predicted advanced AMD in AFR and ASN. First, we explored the 52 variants regarding association and frequency: when comparing results from a logistic regression model containing all 52 variants in EUR-only IAMDGC 2.0 versus IAMDGC 1.0, conditional ORs, standard errors and RAFs were very similar (**Figure 4A-C, Supplementary Data 4**). Ancestry-specific RAFs were, for some variants, substantially diverging between ancestry, which was not due to statistical sampling error (i.e. non-overlapping 95%-CIs; **Supplementary Figure 4A-C**) and thus in line with ancestry-differential genetic architecture. Second, we generated the 52-variant GRS in this IAMDGC 2.0 dataset with the weights from Fritsche et al. (**Methods**). The GRS clearly differentiated cases and controls in the cross-ancestry data (**Figure 4D**). In EUR individuals, the GRS was associated with advanced AMD at P<0.001 and predicted advanced AMD (OR=1.46, AUC=0.80; **Table 2**), as expected since the variants were identified in EUR individuals from IAMDGC 1.0. Importantly, the GRS was also associated with advanced AMD in other ancestries (P<0.001; ORs=1.21 to 1.34 per average risk allele) and predicted advanced AMD in AFR, ASN and Other (AUC=0.65, 0.80, 0.78, respectively; **Table 2**). This pattern was similar with a 42-variant GRS focusing on variants analyzable in each ancestry (risk-increasing allele count, ancestry-specific RAC≥20 and 2^*^n–RAC ≥20; 42-variant GRS; **Table 2**). A GRS focusing on the three of the eight independent *CFH* variants analyzable in each ancestry showed no significant association in AFR but in all other ancestries (OR [95%-CI] =1.11 [0.90, 1.36]). However, the GRS excluding the *CFH* variants (44-variant GRS) performed similar as the 52-variant GRS in AFR and better in all other ancestries (**Table 2**). Thus, the 52-variant GRS performs best to predict advanced AMD across ancestries.

**Figure 4:**
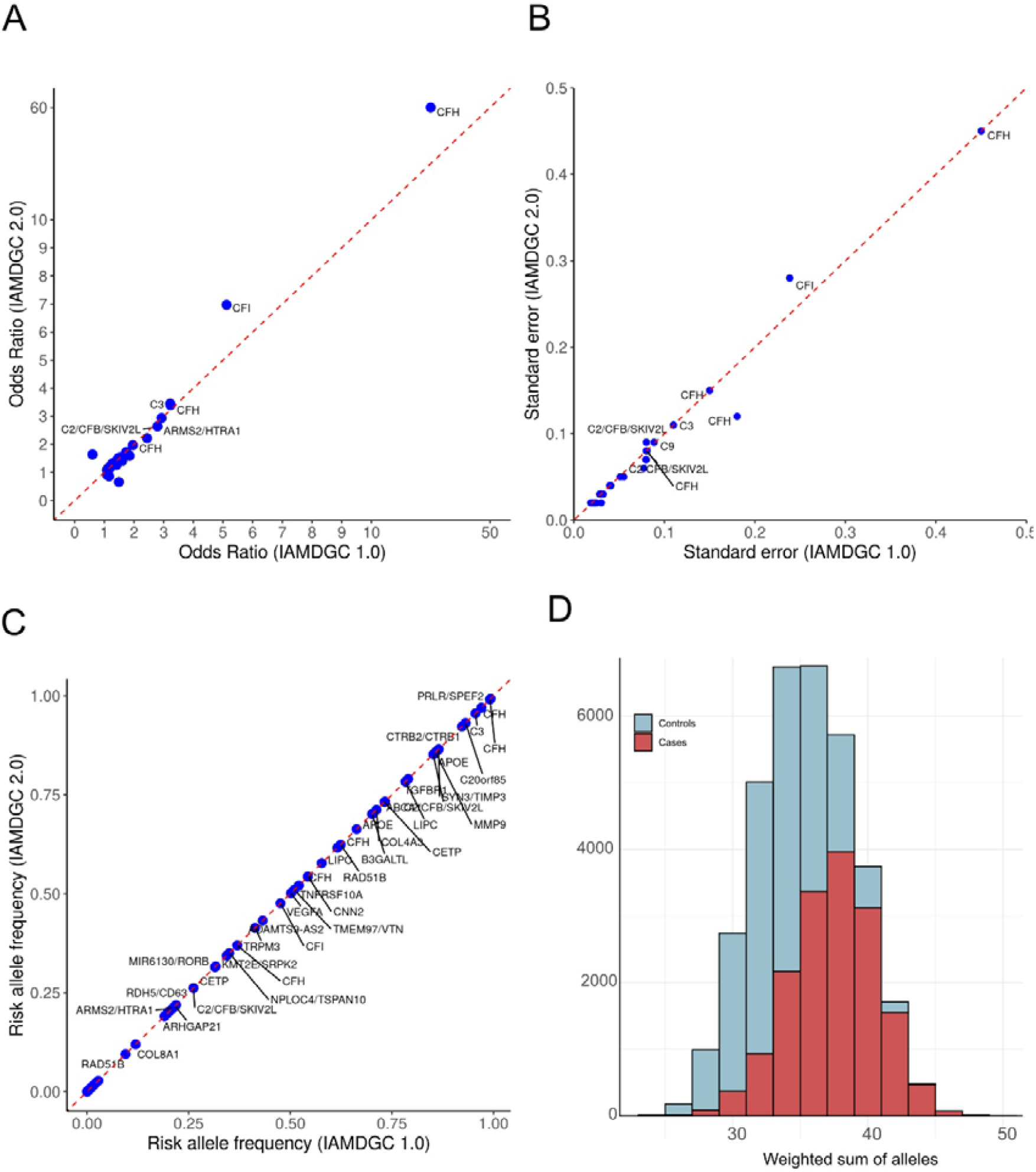
Revisiting the 52 variants identified previously and the 52-variant GRS here in IAMDGC 2.0 cross-ancestry analysis. Previously, 52 independent variants were identified for association with advanced AMD risk and their joint effect is a known GRS for advanced AMD. We derived their association conditional on each other in the IAMDGC 2.0 EUR-only dataset (logistic regression adjusted for ancestry-specific PCs, all 52 variants in one model) and compared their association from IAMDGC 1.0 (EUR-only, **Supplementary Data 4**). Shown are (**A**) conditional odds ratios, (**B**) the respective standard errors (SE) and (**C**) the frequency of the risk-increasing allele (RAF) derived from IAMDGC 1.0 analysis (**X-axis**) compared to the IAMDGC 2.0 EUR-only (**Y-axis**). We derived the GRS with weights from Fritsche et al.^13^ counting risk-increasing alleles in IAMDGC 2.0 (divided by average weight) and show (**D**) the distribution of the 52-variant GRS in individuals across ancestries (cases in red and controls in blue).

We here provide IAMDGC 2.0 as updated data repository with denser genomic coverage. Our cross-ancestry GWAS identified two AMD loci that were new compared to IAMDGC 1.0, which more firmly establish the *CPN1* locus in cross-ancestry data compared to Han et al.^3^ and *SERPINA1* in data of both sexes^4^. Compared to other work using MVP or UK Biobank^4^, the IAMDGC dataset is distinguished by the ophthalmological ascertainment not only for advanced AMD cases, but also for AMD-free controls. This contrasts to diagnostic records for cases and a lack of it for controls (MVP) or self-reported AMD yes or no (UK Biobank as used by Gorman et al.). IAMDGC cases were ascertained for clear signs of advanced AMD differentiated from early or intermediate AMD, while AMD diagnostic records might include such earlier AMD stages, and UK Biobank AMD self-report has been shown to be subject to misclassification^24^. IAMDGC controls were ascertained for having no AMD, while individuals without AMD diagnosis might include individuals with early signs of AMD, which has been shown to deflate AMD association estimates^25^. Generally, misclassification in cases and/or controls is known to bias OR estimates towards unity^26^ and the larger ORs for most AMD variants in IAMDGC (1.0 or 2.0) versus observed ORs by Gorman et al. might be explained, at least in part, by less misclassification in cases and controls. It was particularly important to bridge the gap between EUR-centered GWAS to cross-ancestry analyses. While we need to acknowledge limited sample sizes for AFR and ASN, our ancestry-comparative results on the top two AMD loci suggest no or lower *CFH* risk in AFR compared to EUR in line with other work^4^. Our conclusion for the *HTRA1/ARMS2* locus was different from Gorman et al., as our data and analyses suggest an ancestry-shared signal around *ARMS2* with similar genetic risk across ancestries accompanied by a second rare-variant-signal residing in *HTRA1*. Importantly, our results validate the 52-variant GRS to predict advanced AMD in AFR and ASN and thus establish this GRS for prediction cross-ancestries.

## METHODS

### Ethics Statement

All data was collected according to Declaration of Helsinki principles. Study participants provided informed consent, and protocols were reviewed and approved by local ethics committees.

### Dataset, reference panel, and method for imputation

The IAMDGC genomic data was genotyped and quality controlled centrally as described previously^13^. The same variant selection before imputation was applied here as previously (i.e. genotype call rate <0.985, HWE P<1×10^−6^, or mapping to multiple locations). We lifted the genomic data from build 37 to build 38 for imputation, including full mapping to the build for the TOPMed Michigan Imputation Server. Of the 52,189 individuals used previously for imputation in IAMDGC 1.0, we excluded individuals from Beaver Dam Eye Study (study termination) and individuals with whole genome amplified (WGA) DNA (shown to potentially comprise imputation quality^14^). This resulted in 46,401 individuals included in the IAMDGC 2.0 imputation using TOPMed r2 as reference panel and server (as of January 2023, BioCatalyst imputation server^27^). This TOPMed r2 reference panel consisted of 97,256 reference individuals and 308,107,085 genetic variants distributed across 22 autosomes and the X chromosome. To accommodate maximum sample size requirements of the TOPMed imputation server, we split the IAMDGC data into 2 sets of 23,200 and 23,201 individuals for two imputation batches and merged the data post imputation. The TOPMed imputation server phased genomic data using Eagle v2.4 and imputed untyped variants using minimac4. We evaluated MAF and RSQ from each of the two imputation batches and used mean MAF and mean RSQ for further references of MAF and RSQ.

After imputation, we derived the imputed variant yield by categories of RSQ and MAF and defined well-imputed variants as done previously^13^ (i.e. RSQ≥ 0.3 for MAF≥1%; RSQ≥0.8 for MAF<1%). IAMDGC 2.0 refers to this TOPMed-imputed dataset.

### Ancestry-specific GWAS in IAMDGC 2.0

An individual’s ancestry was defined as before^13^ using data from the Human Genome Diversity Project (EUR, ASN, AFR, and unspecified other ancestry, OTH). Ten ancestry-specific PCs were generated using the SNPRelate R-package^28^ with independent (ld-threshold=0.5) autosomal TOPMed variants shared with Human Genome Diversity Project genotypes.

For ancestry-specific GWAS, we utilized regenie^16^. Regenie conducts ridge regression based on directly genotyped variants to account for relatedness and then, for each genetic variant, derives association of allele dosages with case-control status using logistic ridge regression. We applied regenie using the Firth test option for each well-imputed variant that had a per-ancestry MAC≥20. Such a MAC cut-off is necessary to yield stable statistics for Firth test in balanced case-control comparisons according to Ma et al.^15^. GWAS were adjusted for 2 PCs followed by sensitivity analyses for identified variants adjusting additionally for sex and age or 10 PCs as done previously^13^, and for adjusting for imputation batch. Per ancestry, genomic control lambda was computed based on all analyzed variants and used for GC-correction if lambda>1.0.

### Cross-ancestry meta-analysis GWAS

Ancestry-specific GWAS results were meta-analyzed using the inverse-variance weighted fixed-effect method implement in METAL. Of note, the fixed-effect method is valid for meta-analyses even when ancestry specific effects are not the same; the meta-analyzed effect size reflects the average effect size across ancestries. We computed the lambda of the meta-analyzed variants and applied GC-correction, again, if lambda>1.0.

### Locus identification

First, we screened meta-analyzed GWAS results focused on genetic variants with MAF≥ 1%, since AFR or ASN individuals did not contribute to variants with MAF<1% due to the ancestry-specific MAC≥20 requirement. For this, we judged variants as genome-wide significant if P< 5×10^−8^. For locus identification, we selected the variant with the smallest P-value genome-wide, extracted the locus region (lead variant ±500kb), and then searched the remaining genome, until no further variant reached P<5×10^−8^. We merged overlapping regions. We extended this search to also include rare variants, acknowledging that most rare variants were analyzable only in EUR, and judged variants at P<1×10^−9^ to account for the increased multiple testing burden.

### Comparison of per-ancestry association and per-ancestry allele frequency for the loci near *HTRA1/ARMS2* and *CFH*

We compared per-ancestry ORs by deriving 95%-CIs (based on standard error from Firth test logistic regression adjusted for 2 PCs) and evaluating whether 95%-CIs overlapped across ancestries (no difference, at P<0.05) and with unity (no association, at P<0.05). We compared per-ancestry AF (for risk-increasing alleles) by deriving the standard error via SQRT((AF)^*^(1-AF)/2n) and evaluating the overlap of 95%-CIs. Heterogeneity between ancestry-specific ORs across multiple ancestries was tested by Q-test and quantified by I^229^.

### Statistical fine-mapping using SuSiE and MESuSiE

Based on the well-imputed (RSQ≥ 0.3 for MAF≥ 1%; RSQ≥0.8 for MAF<1%) genetic variants that were analyzable in both EUR and AFR (MAC≥20 per ancestry), we applied MESuSiE for multi-ancestry fine-mapping^19^. In contrast to the default MESuSiE implementation, we kept A/T and C/G SNPs and indels. Since MESuSiE requires sufficient power in each ancestry, we focused on the two top AMD loci. Based on a minimal absolute correlation threshold of 0.5, 95% credible sets were constructed. MESuSiE computes, for each genetic variant, both ancestry-specific posterior inclusion probabilities (PIPs) and shared-ancestry PIPs. For each signal, a posterior probability for the signal to be of shared, EUR-specific, or AFR-specific was computed based on summing up variant-specific PIPs. We also applied SuSiE for single-ancestry fine-mapping in EUR^20^ to compare results. Setting the maximal number of signals per locus to 10 (L=10), the method was executed as implemented in the MESuSiE and susieR R-packages.

### GRS analyses

The GRS was generated by using the individual’s allele dosage for each variant (risk-increasing allele as per Fritsche et al.^13^, multiplying it by the variant’s weight (i.e. beta-estimates from Fritsche et al.), and then deriving the sum across all variants. The GRS was then scaled by dividing through the average weight (sum of weights divided by number of variants in the score), which results in a GRS unit to reflect one risk allele of average effect. We calculated the GRS based on four sets of variants: (i) comprising all 52 variants identified previously^13^ (52-variant-GRS), (ii) using variants outside the *CFH* locus (44-variant-GRS), (ii) including only variants usable to sum up alleles in each ancestry (i.e. RAC ≥20 and 2^*^n – RAC ≥20 in EUR, AFR, ASN, and Other; 42-variant GRS), (iii) using only variants within the *CFH* locus that usable to sum up alleles per ancestry. Based on each of these variant sets, we derived the ORs per ancestry and judged ancestry-specific 95%-CIs for overlapping across ancestries (no difference) or with unity (no association). To compare the performance of ancestry-specific GRS regarding prediction of advanced AMD, we derived the area under the receiver-operating-characteristics (ROC) curve (AUC). Predicted probabilities used for ROC analysis were derived from logistic regression analyses conducted per ancestry (advanced AMD ∼ GRS + PC1 + PC2).

## Data availability

GWAS summary statistics will be made available (upon publication) on the websites of the IAMDGC data holders in the United States (Cleveland; http://amdgenetics.org/) and Germany (Regensburg; https://www.genepi-regensburg.de/gwas-summary-statistics). Data permitted for sharing by respective institutional review boards have been deposited in the database of Genotypes and Phenotypes (dbGaP) under accession phs001039.v1.p1.

## Code availability

Genetic data were analyzed using regenie (v3.2.8) available at https://rgcgithub.github.io/regenie/. Meta-analyses were performed using METAL (version released on 2018-02-12) available at https://csg.sph.umich.edu/abecasis/metal/download/. Genotypes were imputed on the BioData CATALYST TOPMed Imputation Server (https://imputation.biodatacatalyst.nhlbi.nih.gov/). Manhattan and QQ-Plots were generated with EasyQC available at https://www.genepi-regensburg.de/easyqc. Regional association plots were generated with Locuszoom (v1.4) (http://locuszoom.org/). Genetic Risk Score analyses were performed with R (v4.4.2) available at https://www.r-project.org/.

## Acknowledgements

This work was supported by NIH grant R01 EY022310 (to J.L.H. and M.A.V.), NIHG RES516564 (to I.M.H), BrightFocus Fellowship for Macular Degeneration M2021006F (to M.Gr.) and NIH 1X01HG006934-01 for CIDR. Further group-specific acknowledgements are stated in the **Supplementary Material**.

## Author contributions

I.M.H. and J.L.H conceived the design of analyses, contributed analyses tools, and supervised all work. M.Go. and M.Gr. conducted statistical analysis and interpreted results together with. I.M.H., J.L.H. All authors contributed to manuscript writing (I.M.H., J.L.H., M.Go, M.Gr, J.M.H., B.F., M.B., N.W., W.S.B., Y.E.S., X.Z., S.H.B. and M.A.V.).

