## Supplementary Material for "Diverse ancestry GWAS for advanced age-related macular degeneration in TOPMed-imputed and Ophthalmologically-confirmed 16,108 cases and 18,038 controls"

for


| | |
|---|---|
| **Supplementary Table 1** | Frequency of TOPMed-imputed variants in IAMDGC 2.0. |
| **Supplementary Table 2** | Results from cross-ancestry GWAS in IAMDGC 2.0 for 36 AMD locus lead variants. |
| **Supplementary Figure 1** | Manhattan Plots of ancestry-specific GWAS in IAMDGC 2.0. |
| **Supplementary Figure 2** | Association signals of seven loci with moderate correlation between the IAMDGC 2.0 and the IAMDGC 1.0 lead variants. |
| **Supplementary Figure 3** | Comparison of the 36 locus lead variants from the IAMDGC 2.0 GWAS regarding association and frequency in IAMDGC 2.0 versus IAMDGC 1.0. |
| **Supplementary Figure 4** | Revisiting the 52 variants identified previously in IAMDGC 2.0 cross-ancestry analysis. |

**Group Specific Acknowledgements**

**Supplementary Table 1: Frequency of TOPMed-imputed variants in IAMDGC 2.0.**

We imputed the 46,601 individuals with measured genotypes with the TOPMed references data (Biocatalyst imputation server) and obtained more than 292 million autosomal variants. We here show absolute numbers and relative frequencies of these variants by Minor Allele Frequency (MAF) and imputation quality (RSQ) categories.

| MAF category | RSQ category | #Variants (%) |
|---|---|---|
| Total | Low | 158,951,074 (54.43%) |
| | Medium | 91,297,455 (31.26%) |
| | High | 41,779,192 (14.31%) |
| Monomorphic MAC=0 | Low | 97,083,508 (33.24%) |
| | Medium | 0 (0%) |
| | High | 0 (0%) |
| Rare MAF<1% | Low | 61,707,532 (21.13%) |
| | Medium | 90,565,414 (31.01%) |
| | High | 33,489,893 (11.47%) |
| Less frequent MAF 1%-5% | Low | 137,680 (0.05%) |
| | Medium | 436,516 (0.15%) |
| | High | 2,315,925 (0.79%) |
| Common MAF>5% | Low | 22,354 (0.01%) |
| | Medium | 295,525 (0.1%) |
| | High | 5,973,374 (2.05%) |
| Total | | 292,027,721 (100%) |

Shown are absolute and relative frequencies of autosomal variants by RSQ categories (low: RSQ<0.3, medium: 0.3≤RSQ<0.8, high: 0.8≤RSQ) and by MAF categories (monomorphic: MAF=0, rare: 0<MAF<0.01, less frequent: 0.01≤MAF≤0.05, common: 0.05<MAF).

**Supplementary Table 2: Results from cross-ancestry GWAS in IAMDGC 2.0 for 36 AMD locus lead variants.**

We conducted cross-ancestry GWAS for advanced AMD (16,108 advanced AMD cases, 18,038 AMD-free controls; Firth test based logistic regression adjusted for ancestry-specific PCs, using regenie). Shown are association statistics for 36 AMD locus lead variants: (i) the 26 among the 34 established AMD loci identified in IAMDGC 2.0 at $P<5x10^{-8}$, the two additional loci discovered at $P<5x10^{-8}$, and the lead variants of the other 8 of the 34 known AMD locus identified at $P<1x10^{-4}$ in the IAMDGC 2.0 cross-ancestry GWAS. Also shown is the correlation of these lead variants with the lead variants identified previously[1] (IAMDGC 1.0). Results from ancestry-specific GWAS in IAMDGC 2.0 for these lead variants as well as the lead variants identified in IAMDGC 1.0 are shown in **Supplementary Data 1.** Region plots of loci where the lead variants did not correlate (r<0.8) are shown in **Supplementary Figure 2**.

| Locus Name | RSid | Chr | Pos | Rsq | RA | OA | RAF | cross-ancestry analysis IAMDGC 2.0 | | | R with IAMDGC 1.0 lead variant |
|---|---|---|---|---|---|---|---|---|---|---|---|
| | | | | | | | | OR | SE | P | |
| **The 26 loci among the 34 loci validated with genome-wide significance** | | | | | | | | | | | |
| *CFH* | rs3766405 | 1 | 196,726,031 | 1.00 | C | T | 0.67 | 2.60 | 0.02 | $1.63x10^{-466}$ | 1.00 |
| *ADAMTS9-AS2* | rs17727064 | 3 | 64,728,506 | 1.00 | G | A | 0.46 | 1.14 | 0.02 | $2.09x10^{-11}$ | 1.00 |
| *COL8A1* | rs116675086 | 3 | 99,527,075 | 0.99 | A | T | 0.018 | 1.67 | 0.07 | $7.76x10^{-13}$ | 0.96 |
| *CFI* | rs13112432 | 4 | 109,660,831 | 0.99 | A | T | 0.70 | 1.17 | 0.02 | $2.04x10^{-14}$ | 0.74 |
| *C9* | rs62358364 | 5 | 39,345,204 | 0.99 | T | G | 0.013 | 1.70 | 0.09 | $1.74x10^{-09}$ | 0.94 |
| *C2-CFB-SKIV2L* | rs556679 | 6 | 31,926,578 | 1.00 | C | T | 0.91 | 1.92 | 0.04 | $2.34x10^{-77}$ | 0.89 |
| *VEGFA* | rs943080 | 6 | 43,858,890 | 1.00 | T | C | 0.53 | 1.14 | 0.02 | $8.71x10^{-12}$ | 1.00 |
| *PILRB-PILRA* | rs75636500 | 7 | 100,350,034 | 1.00 | G | C | 0.19 | 1.15 | 0.02 | $3.72x10^{-09}$ | 0.97 |
| *KMT2E-SRPK2* | rs569309787 | 7 | 105,079,829 | 0.97 | C | CATT | 0.49 | 1.13 | 0.02 | $1.78x10^{-10}$ | 0.74 |
| *TNFRSF10A* | rs13278062 | 8 | 23,225,458 | 0.89 | T | G | 0.53 | 1.13 | 0.02 | $5.89x10^{-10}$ | 1.00 |
| *TGFBR1* | rs1626340 | 9 | 99,161,090 | 1.00 | G | A | 0.79 | 1.15 | 0.02 | $2.04x10^{-09}$ | 1.00 |
| *ARHGAP21* | rs72784286 | 10 | 24,639,330 | 0.98 | T | C | 0.21 | 1.14 | 0.02 | $3.31x10^{-08}$ | 0.97 |
| *ARMS2-HTRA1* | rs61871747 | 10 | 122,453,530 | 1.00 | T | C | 0.32 | 2.88 | 0.02 | $1.68x10^{-581}$ | 1.00 |
| *RDH5-CD63* | rs3138142 | 12 | 55,721,801 | 0.84 | T | C | 0.23 | 1.19 | 0.02 | $2.14x10^{-12}$ | 1.00 |
| *ACAD10* | rs7296313 | 12 | 111,440,722 | 0.99 | T | C | 0.78 | 1.14 | 0.02 | $3.72x10^{-08}$ | 0.06 |
| *RAD51B* | rs12883719 | 14 | 68,343,500 | 0.99 | T | G | 0.63 | 1.11 | 0.02 | $4.86x10^{-08}$ | 0.98 |

| | | | | | | | | | | | |
|---|---|---|---|---|---|---|---|---|---|---|---|
| *LIPC* | rs2043085 | 15 | 58,388,755 | 0.87 | C | T | 0.62 | 1.16 | 0.02 | $1.70x10^{-13}$ | 1.00 |
| *CETP* | rs5817082 | 16 | 56,963,437 | 0.99 | C | CA | 0.74 | 1.19 | 0.02 | $5.13x10^{-16}$ | 1.00 |
| *CTRB2-CTRB1* | rs9936550 | 16 | 75,208,952 | 0.98 | C | T | 0.93 | 1.27 | 0.04 | $2.57x10^{-10}$ | 0.94 |
| *NPLOC4-TSPAN10* | rs34635363 | 17 | 81,582,224 | 0.96 | A | G | 0.36 | 1.13 | 0.02 | $1.95x10^{-09}$ | 0.96 |
| *C3* | rs2230199 | 19 | 6,718,376 | 1.00 | C | G | 0.23 | 1.46 | 0.02 | $1.86x10^{-63}$ | 1.00 |
| *APOE* | rs429358 | 19 | 44,908,684 | 0.99 | T | C | 0.88 | 1.49 | 0.03 | $2.45x10^{-40}$ | 1.00 |
| *MMP9* | rs6065912 | 20 | 46,003,728 | 0.99 | G | A | 0.88 | 1.19 | 0.03 | $1.32x10^{-09}$ | 0.96 |
| *C20orf85* | rs57619911 | 20 | 58,070,144 | 0.98 | G | T | 0.89 | 1.22 | 0.03 | $6.46x10^{-11}$ | 0.84 |
| *SYN3-TIMP3* | rs7289865 | 22 | 32,695,826 | 1.00 | A | C | 0.85 | 1.27 | 0.03 | $7.08x10^{-18}$ | 0.98 |
| *SLC16A8* | rs11089861 | 22 | 38,106,818 | 0.99 | G | C | 0.22 | 1.15 | 0.02 | $9.55x10^{-10}$ | 0.92 |
| **Two loci identified with genome-wide significance not among the 34 IAMDGC 1.0 loci** | | | | | | | | | | | |
| *CPN1* | rs17112478 | 10 | 100,045,603 | 0.99 | T | A | 0.96 | 1.29 | 0.04 | $8.32x10^{-09}$ | 1.00 |
| *SERPINA1* | rs28929474 | 14 | 94,378,610 | 1.00 | C | T | 0.98 | 1.54 | 0.08 | $2.95x10^{-08}$ | 1.00 |
| **The eight loci among the 34 IAMDGC 1.0 loci identified at $P<1x10^{-4}$** | | | | | | | | | | | |
| *COL4A3* | rs11674609 | 2 | 227,176,229 | 0.91 | T | C | 0.29 | 1.09 | 0.02 | $4.37x10^{-05}$ | 0.01 |
| *PRLR-SPEF2* | rs17510915 | 5 | 35,460,175 | 0.95 | G | A | 0.93 | 1.17 | 0.04 | $3.31x10^{-05}$ | 0.58 |
| *TRPM3* | rs188092309 | 9 | 70,896,789 | 0.97 | A | C | 0.03 | 1.38 | 0.06 | $5.13x10^{-08}$ | 0.21 |
| *MIR6130-RORB* | rs10781171 | 9 | 73,950,568 | 1.00 | C | A | 0.33 | 1.11 | 0.02 | $8.13x10^{-07}$ | 0.99 |
| *ABCA1* | rs2740488 | 9 | 104,899,461 | 0.97 | A | C | 0.74 | 1.09 | 0.02 | $5.01x10^{-05}$ | 1.00 |
| *B3GALTL* | rs7981280 | 13 | 31,245,254 | 0.99 | C | T | 0.70 | 1.11 | 0.02 | $9.12x10^{-07}$ | 1.00 |
| *TMEM97-VTN* | rs708100 | 17 | 28,361,641 | 0.98 | A | G | 0.53 | 1.09 | 0.02 | $1.15x10^{-05}$ | 0.98 |
| *CNN2* | rs3752241 | 19 | 1,053,525 | 0.95 | G | C | 0.17 | 1.12 | 0.03 | $4.27x10^{-06}$ | 0.34 |

RSid=rs-number, Chr=Chromosome, Pos=Position on GRCh38, Rsq=Imputation quality, RA=Risk Allele, OA=Other Allele, RAF=Risk Allele Frequency, OR=Odds Ratio, SE=Standard error, P=P-value, R=Spearman correlation R.

**Supplementary Figure 1. Manhattan Plots of ancestry-specific GWAS in IAMDGC 2.0.**

We conducted GWAS in European (EUR), African (AFR), Asian (ASN) and in any other ancestry (OTH) for advanced AMD in the IAMDGC 2.0 data ($n_{cases}/n_{controls}$ = 15,616/16,723, 50/357, 207/322, 235/636, respectively). GWAS of EUR and OTH ancestries were GC-corrected (GC-lambda: 1.11 and 1.08, respectively). GWAS of AFR and ASN were not corrected for genomic inflation (GC-lambda: 0.99 and 0.98, respectively). We here present Manhattan Plots for (**A**) EUR, (**B**) AFR, (**C**) ASN and (**D**) other ancestry. We color-marked the 26 among the 34 loci known AMD loci[1] in green, the two new loci in red and the other 8 of the 34 known AMD loci in blue.

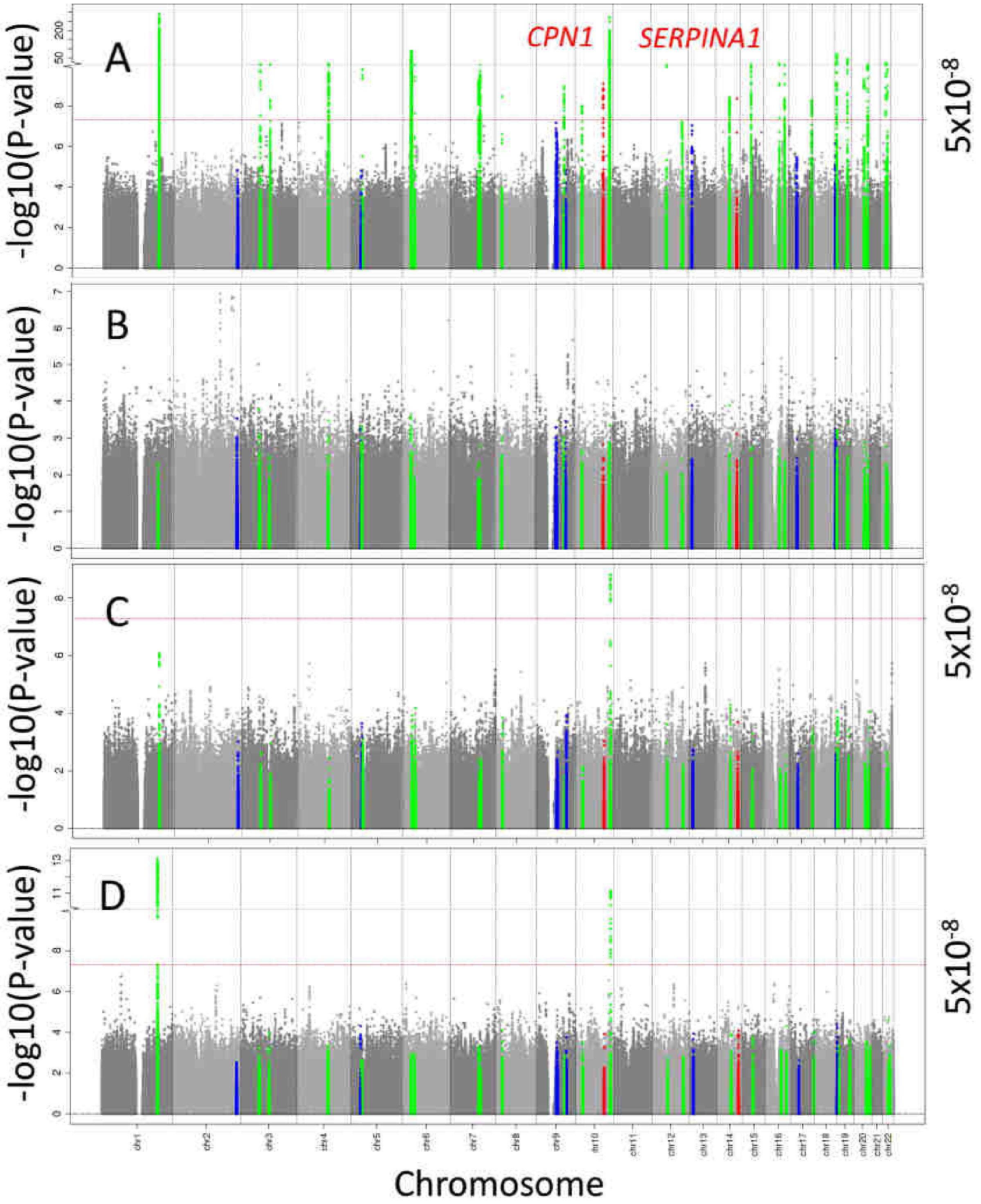

**Supplementary Figure 2. Association signals of seven loci with moderate correlation between the IAMDGC 2.0 and the IAMDGC 1.0 lead variants.**

We mapped the genome-wide significant AMD loci identified in the cross-ancestry IAMDGC 2.0 analysis to the previously established 34 loci in the IAMDGC 1.0[1] and found seven loci with moderately correlated locus lead variants (r<0.8). We here present the regional association signals around (**A&B**) *CFI*, (**C&D**) *KMT2E-SRPK2* (**E&F**) *ACAD10*, (**G&H**) *COL4A3* (**I&J**) *PRLR-SPEF2*, (**K&L**) *TRPM3* and (**M&N**) *CNN2* loci. We present the association with the minus log10 P-value (**Y-axis**) versus genomic position (**X-axis**) in the IAMDGC 2.0 (left column) and in the IAMDGC 1.0 (right column) data. We indexed the locus lead variant by a violet diamond and added the rs-number of the lead variant of the other data set, respectively.

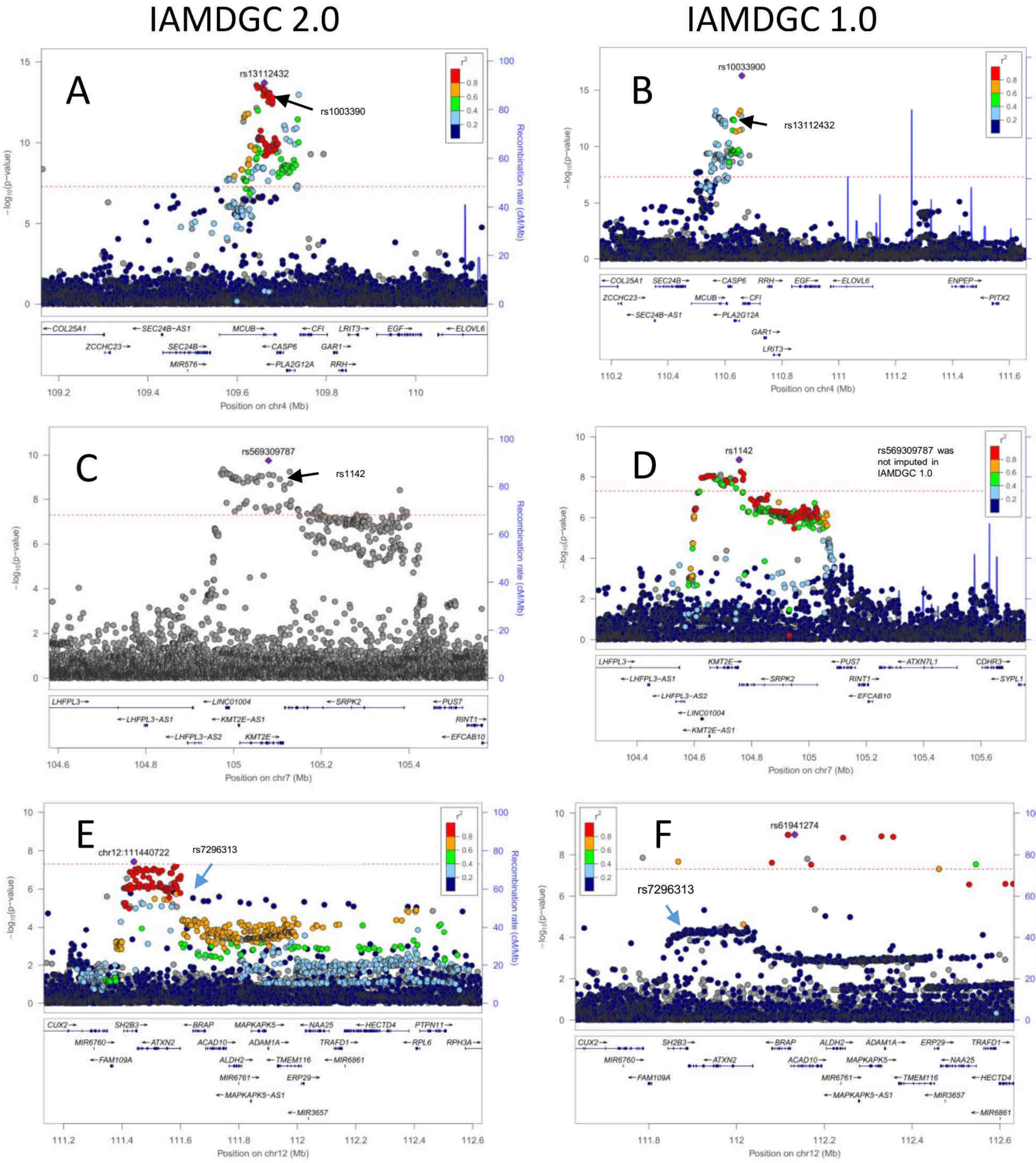

**Supplementary Figure 2 continued.**

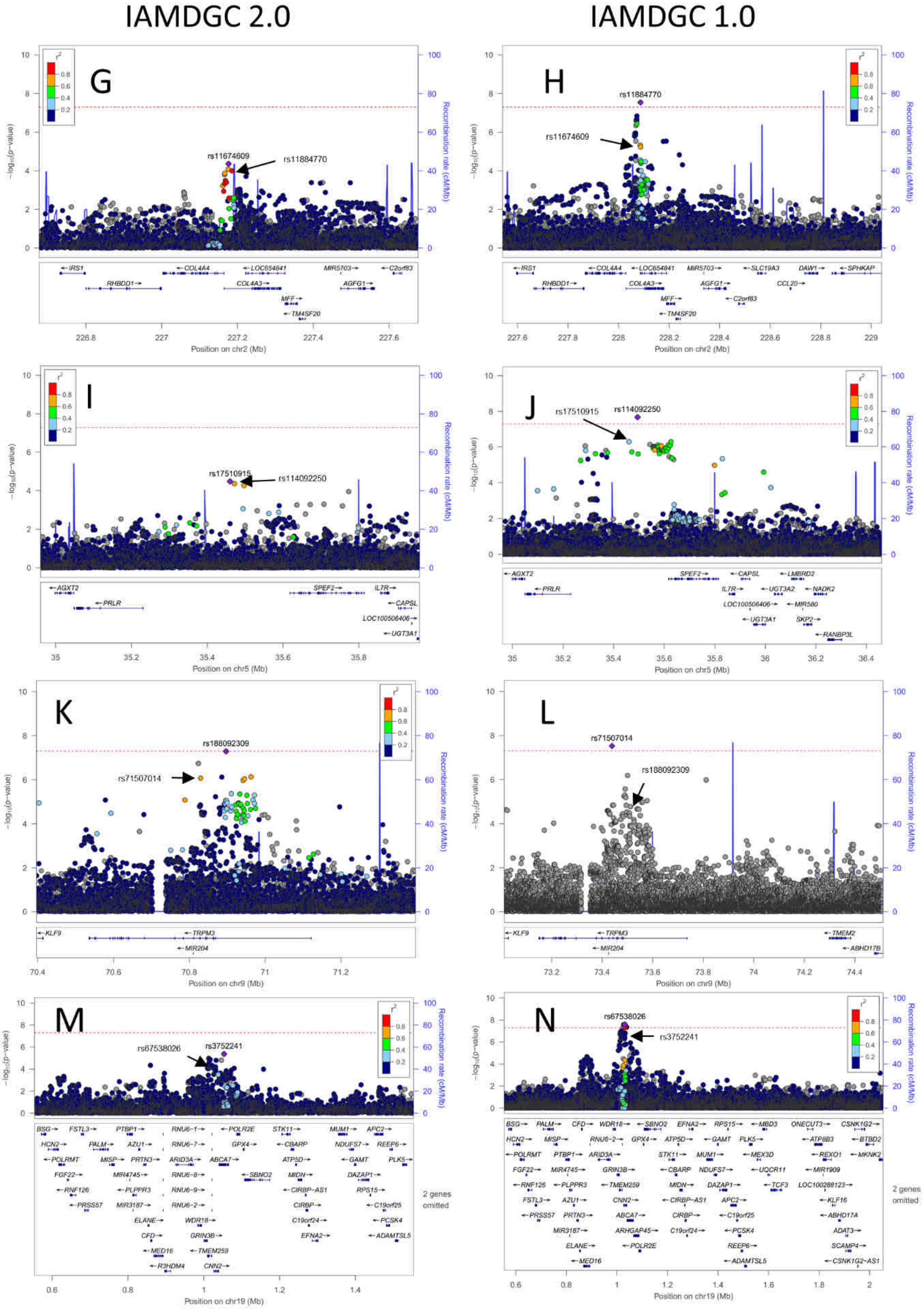

**Supplementary Figure 3. Comparison of the 36 locus lead variants from the IAMDGC 2.0 GWAS regarding association and frequency in IAMDGC 2.0 versus IAMDGC 1.0.**

We identified 34 locus lead variants of previously identified AMD risk loci[1] and the two newly identified loci in the IAMDGC 2.0 (*SERPINA1* and *CPS1*) in the cross-ancestry GWAS of 16,108 late AMD cases and 18,038 AMD-free controls. We evaluated, if the identified locus lead variants' association and frequencies were comparable between IAMDGC 2.0 cross-ancestry versus IAMDGC 2.0 EUR-only, and between IAMDGC 2.0 EUR-only versus IAMDGC 1.0. We show (**A&B**) Odds Ratios, (**C&D**) standard errors and (**E&F**) risk allele frequency (i.e. AMD risk-increasing allele in cross-ancestry IAMDGC 2.0). We illustrate (left) IAMDGC 2.0 cross-ancestry versus IAMDGC 2.0 EUR-only results and (right) IAMDGC 2.0 EUR-only versus IAMDGC 1.0.

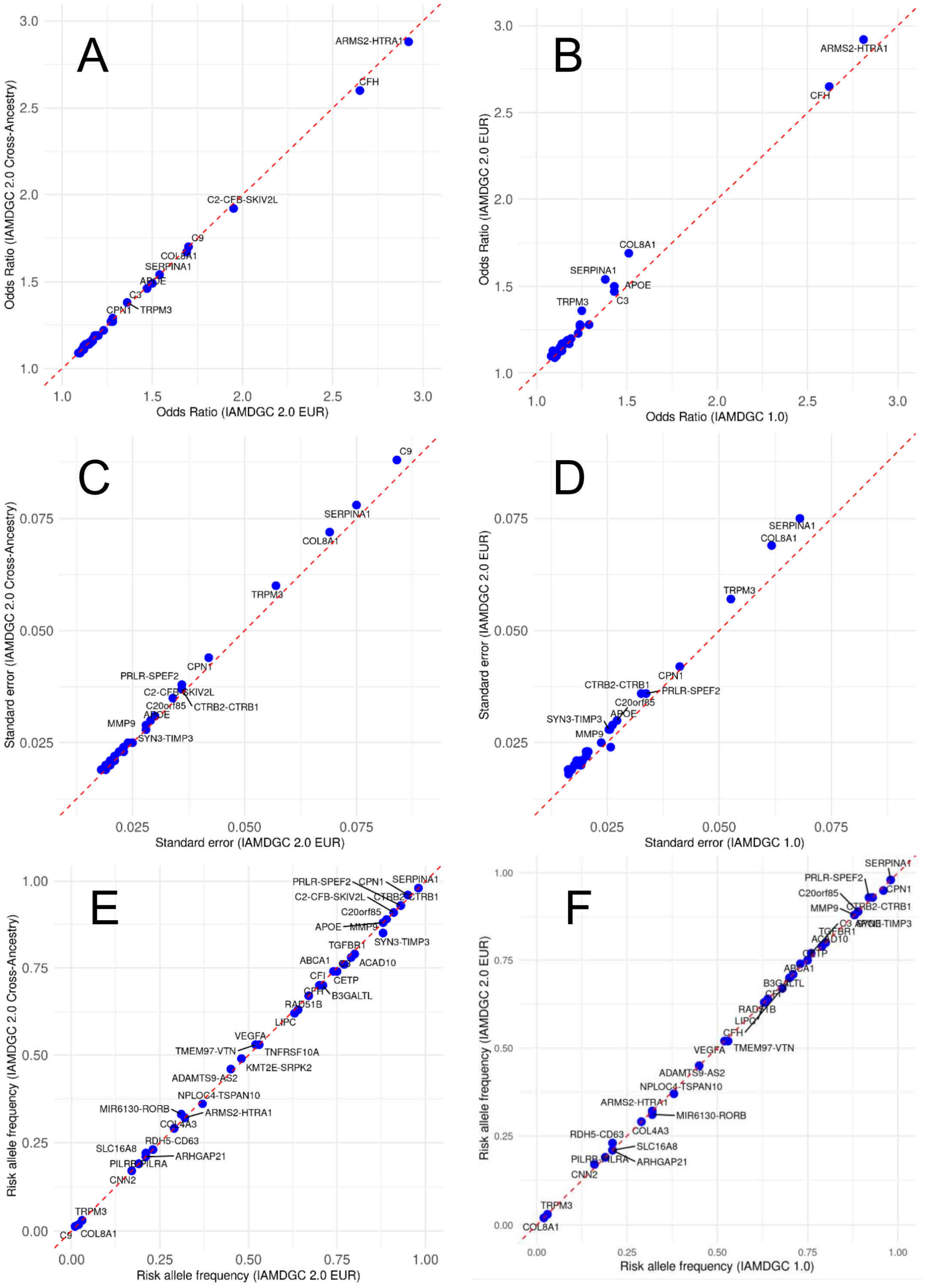

**Supplementary Figure 4: Revisiting the 52 variants identified previously in IAMDGC 2.0 cross-ancestry analysis.**

Previously, 52 independent variants were identified for association with advanced AMD risk and their joint effect is a known GRS for advanced AMD. We derived their association conditional on each other in the IAMDGC 2.0 EUR-only dataset (logistic regression adjusted for ancestry-specific PCs, all 52 variants in one model) and compared their association from IAMDGC 1.0 (EUR-only, **Supplementary Data 4**). Shown are the frequency of the risk-increasing allele (RAF, **X-axis**) in individuals of (**A**) African (AFR), (**B**) Asian (ASN) and (**C**) Other (OTH) ancestry compared to Europeans (EUR, **Y-axis**). Whiskers in black represent 95% confidence intervals of the RAF (RAF±1.96*SE).

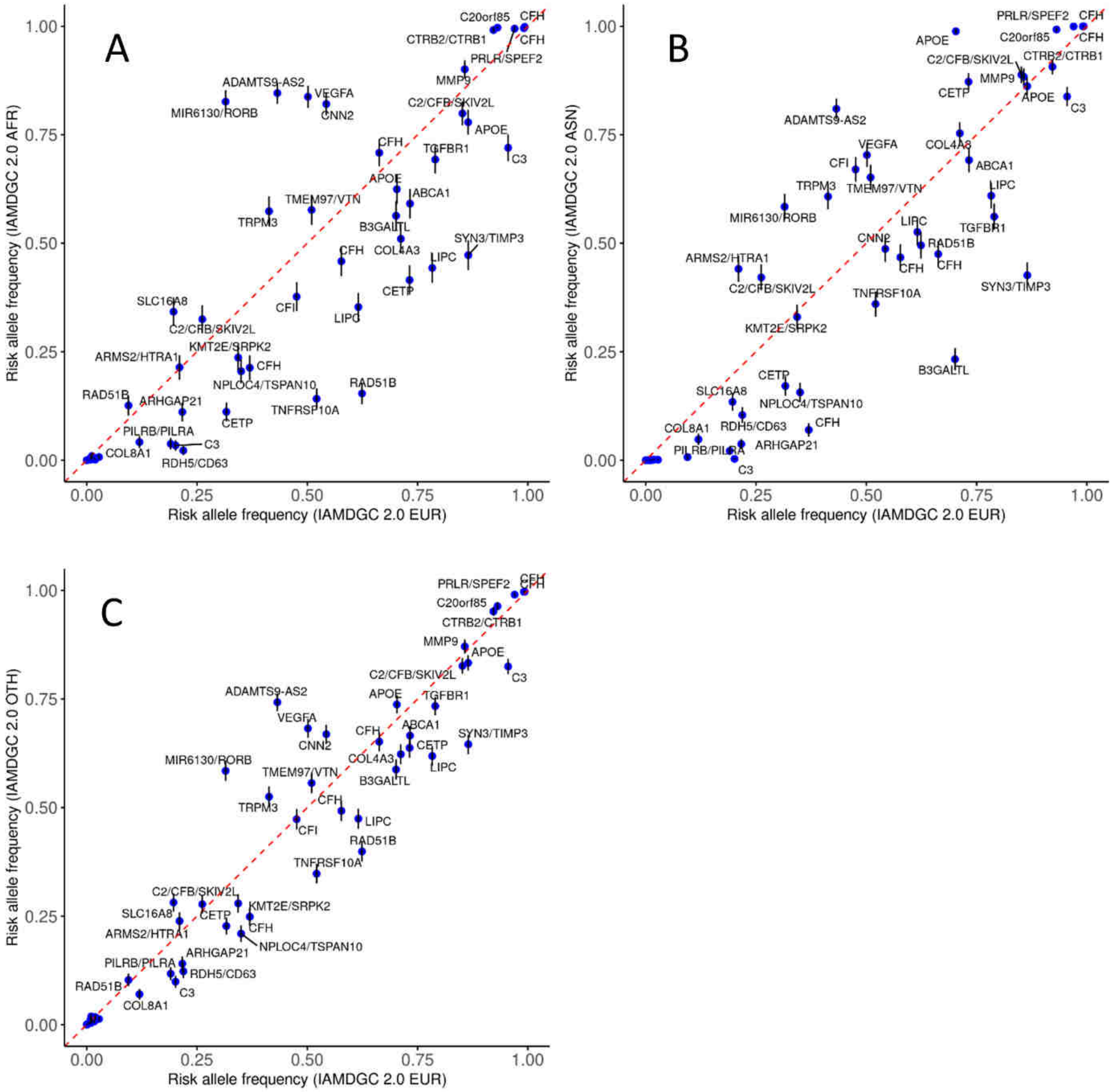

**Group Specific Acknowledgements**

The UWA, LEI & Flinders group acknowledges financial support for participant recruitment and sample processing provided by the National Health and Medical Research Council (NHMRC) of Australia (#1023911), the Ophthalmic Research Institute of Australia, the BrightFocus Foundation and a Ramaciotti Establishment Grant. CERA receives Operational Infrastructure Support from the Victorian Government. KPB, JEC and AWH are supported by NHMRC Fellowships. The authors acknowledge the support of B. Usher-Ridge, L. Palmer, L Ma and DL Lim in patient recruitment and data collection.

The Pittsburgh group acknowledges funding to MBG from NIH/NEI R01 EY09859, Research to Prevent Blindness (N.Y, N.Y.), Harold and Pauline Price Foundation.

The Cambridge group was supported by the Medical Research Council, UK (grant G0000067 to JRWY, ATM), the Macular Disease Society (JRWY, ATM); the Guide Dogs for the Blind Association (ATM, JRWY) and the Department of Health's NIHR Biomedical Research Centre for Ophthalmology at Moorfields Eye Hospital and UCL Institute of Ophthalmology. We thank the clinicians who helped with recruitment, the Reading Centre at Moorfields Eye Hospital, London for grading fundus photographs and the subjects who participated in the research.

The EUGENDA-Cologne group was supported by a grant from the Retinovit foundation.

The Vanderbilt group was supported by the National Institutes of Health Grants AG019085 (JLH), EY023164 (JLH), EY022310 (JLH), EY012118 (JLH, AA, MAB), AG044089 (JDH), T32 EY007157 (JNCB) and a PhRMA Informatics fellowship (JNCB).

The MMAP-Penn group was supported by the Arnold and Mabel Beckman Initiative for Macular Research (CAC), Research to Prevent Blindness Inc (CAC), EyeSight Foundation of Alabama (CAC), and NIH EY023164 (DS).

The Oregon group was supported by National Eye Institute grants EY021532, and EY0105712, and an unrestricted departmental grant from Research to Prevent Blindness.

The Edinburgh group using the Scottish AMD study was funded by the Chief scientists Office (Scotland) CSO reference number: CZB/4/79 and would like to thank Alan Wright, Ana Armbrecht and Fraser Imrie for collecting the samples and all of the individuals who participated in this study.

The EU/JHU study acknowledges the support of the CEPH Biological Resource Centre by the French Ministère de l'Enseignement Supérieur et de la Recherche, Foundation Fighting Blindness Clinical Research Institute (FFB, CRI), an unrestricted grant to the Wilmer Eye Institute from Research to Prevent Blindness, and Baylor-Johns Hopkins Center for Mendelian Genetics (National Human Genome Research Institute, NHGRI/NIH; 1U54HG006542-01).

The Jerusalem study was supported by grants from the Israel Science fund (ISF) and the Israeli Ministry of Health.

The Southampton study acknowledges Southampton Wellcome Trust Clinical Research Facility for research nurse support in collecting DNA samples, Helen Griffiths (Clinical and Experimental Sciences, University of Southampton) for technical support in processing DNA and all the patients who contributed to this work. AJL supported at the University of Southampton by funding from The Wellcome Trust (076169/A), American Health Assistance Foundation (M2007110), Macula Vision Research Foundation, TFC Frost Charitable Trust, Brian Mercer Charitable Trust, Macular Society, Hobart Trust and the Gift of Sight appeal.

The Marshfield group was supported by grants NIH NCATS: UL1TR000427, NIH NHGRI: 1U01HG006389, and support from the Marshfield Clinic Research Foundation.

The Melbourne study was supported by the National Health and Medical Research Council Australia, project grant 1008979, Centre for Clinical Research Excellence #529923 - Translational Clinical Research in Major Eye Diseases. NHMRC Research Fellowship (PNB, #1028444). CERA receives Operational Infrastructure Support from the Victorian Government.

The Miami group was supported by National Institutes of Health Grants R01 EY012118 (MAP-V, WKS, JLK, SGS, MDC), EY023164 (MAP-V, WKS, JLH, MAP-V), EY022310 (MAP-V) and T32 EY023194 (RJS) and P30-EY005722. All Bascom Palmer Eye Institute authors are partially supported by NIH Center Core Grant P30EY014801 and an unrestricted grant from Research to Prevent Blindness, New York, NY, USA.

The MMAP-Michigan and AREDS groups were supported by Intramural Research Program of the National Eye Institute (ZO1 EY000475); the AREDS study was supported by the National Eye Institute/National Institutes of Health, (contract no.: HHS-NOI-EY-0-2127), Bethesda Maryland; the AREDS2 study was supported by the intramural program funds and contracts from the National Eye Institute/National Institutes of Health (NEI/NIH), Department of Health and Human Services, Bethesda, MD. Contract No. HHS-N-260-2005-00007-C. ADB Contract No. N01-EY-5-0007.

The Michigan study was supported by the National Eye Institute (EY0022005) and the National Human Genome Research Institute (HG006513 HG007022), Foundation Fighting Blindness and National Institutes of Health/National Eye Institute Grant-EY016862.

The NHS/HPF studies were supported by EY021900, EY017362, EY13824, EY009611, CA87969, CA49449, and HL35464.

The Regensburg group was supported by BMBF-01ER1206 (to IMH), EFKS 2012_A147 (IMH), BMBF-01GP1308 (IMH), the Deutsche Forschungsgemeinschaft (grant WE 1259/19-1 and WE1259/19-2, BHFW), and the Alcon Research Institute (BHFW). The Regensburg Team would also like to thank Randy Rueckner for technical assistance.

The Rotterdam-Clinic study was supported by ZoNMW project number: 170885606, MDfonds, Landelijke Stichting voor Blinden en Slechtzienden (LSBS).

The UCSD study was supported by grants from NIH (grants EY014428, EY018660, P30EY022589), 863 Program (2014AA021604), and Research to Prevent Blindness. ZS is supported by 863 Program (2014AA021604), ZY is supported by National Natural Science Foundation of China (81170883 and 81430008), KZ is supported by NIH grants (1R01EY018660-01A10) and VA Merit Award.

The EUGENDA-Neijmegen study was supported by MD Fonds, Gelderse Blindenstichting, Algemene Vereniging ter Voorkoming van Blindheid, Stichting Nederlands Oogheelkundig Onderzoek, Oogfonds.

The Utah study was supported by the ALSAM Foundation, an unrestricted grant from Research to Prevent Blindness to the Department of Ophthalmology and Visual Sciences, University of Utah, SOM, Moran Eye Center.

The Seoul National University Bundang group was supported by grants from the National Research Foundation of Korea, funded by the Ministry of Education, Science, and Technology (grant numbers; NRF-2009-0072603 and NRF-2012R1A1A2008943).

The Westmead/Sydney samples were collected in three studies that were supported by the National Health and Medical Research Council (NHMRC), Australia: Grant IDs 974159, 211069, 457349 and 512423 supported the Blue Mountains Eye Study that provided population-based controls; Grant ID 302010 supported the Cataract Surgery and Risk of Age-related Macular Degeneration study that provided clinic-based early and late AMD cases and controls; and Grant ID 571013 supported the Genes and Environment in late AMD study that provided clinic-based late AMD cases. NHMRC Senior Research Fellowship (JJW, 358702, 632909). NHMRC Senior Research Fellowship (JJW, 358702, 632909). The NHMRC had no role in the design or conduct of these studies.

The Columbia study was supported by the National Institutes of Health/NIH grants R01-EY013435 and P30-EY019007, and Research to Prevent Blindness (New York, NY).

The CWRU group was supported by VA Merit Review (NSP), Foundation Fighting Blindness (SAH), Research to Prevent Blindness (SAH); International Retinal Research Foundation (SKI).
